# Optical coherence tomography as a biomarker for frontotemporal dementia: a systematic review & meta-analysis

**DOI:** 10.64898/2026.05.19.26353366

**Authors:** Eric Wang, Anish Kohli, Hash Brown Taha

**Author notes:** **Corresponding Author** Hash Brown Taha. **Authorship contribution:** EW and HBT conceptualized and designed the study. HBT, EW and AK wrote the manuscript. EW and AK collected the data. EW and HBT analyzed the data. HBT supervised the study. **Declarations: Sources of funding:** None. **Competing Interests:** None.

## Abstract

**Background:** Frontotemporal dementia (FTD) lacks widely accessible disease-specific biomarkers. Optical coherence tomography (OCT) and OCT angiography (OCTA) may provide non-invasive measures of retinal changes associated with neurodegeneration. We conducted a systematic review and meta-analysis evaluating retinal biomarkers in FTD compared with Alzheimer disease (AD) and controls.

**Methods:** A systematic search of PubMed and Embase was conducted through April 25, 2026 according to PRISMA guidelines. Studies evaluating OCT/OCTA biomarkers in FTD with comparator groups were included. Inverse weighted random-effects models, publication bias assessments, and meta-regressions were performed.

**Results:** Ten studies involving 139 individuals with FTD, 87 with AD, 29 with mild cognitive impairment, 14 with TDP-43 proteinopathy, 5 with tauopathy, and 255 controls were included in the systematic review; five studies were eligible for meta-analysis. Compared with AD, individuals with FTD demonstrated significantly thinner retinal nerve fiber layer (RNFL) thickness (SMD = −0.61, 95% CI −0.98, −0.24). Compared with controls, individuals with FTD exhibited significantly thinner ganglion cell layer-inner plexiform layer (GCL-IPL) thickness (SMD = −0.55, 95% CI −1.02, −0.08), whereas pooled analyses across multiple retinal biomarkers were non-significant (SMD = −0.19, 95% CI −0.52, 0.14). RNFL thickness correlated negatively with female % in FTD and positively with age in both AD and controls.

**Conclusions:** Individuals with FTD exhibit lower RNFL thickness than AD and lower GCL-IPL thickness than controls, suggesting retinal alterations may reflect neurodegeneration. However, larger longitudinal studies with standardized OCT/OCTA protocols are needed to determine the diagnostic and prognostic utility of retinal biomarkers in FTD

## Introduction

Dementia is a clinical syndrome characterized by progressive decline in cognition, behavior, and functional independence. Among its subtypes, frontotemporal dementia (FTD) is distinguished by early changes in behavior, personality, and language rather than the prominent memory impairment seen in Alzheimer disease (AD). FTD encompasses a spectrum including behavioral variant FTD (bvFTD) and primary progressive aphasia, and typically affects younger individuals compared to AD [1]. Importantly, FTD arises from distinct underlying pathophysiologic mechanisms, most commonly frontotemporal lobar degeneration driven by tau, TDP-43, or FUS proteinopathies, in contrast to the amyloid and tau-based pathology of AD [2].

Differential diagnosis of dementias remains challenging due to symptom overlap. Given these fundamental biological differences, accurate and early diagnosis is needed for appropriate clinical management and enabling timely intervention and advancing the development of disease-modifying therapeutics. FTD lacks widely accessible, disease-specific markers for routine clinical use. Some advances have been made with neurofilament light chain (NfL) as a marker of neurodegeneration and genetic testing in familial cases, but these lack specificity for FTD subtypes or early diagnosis [3]. Neuroimaging findings, such as frontal and temporal lobe atrophy on MRI or hypometabolism on PET, can support diagnosis but are not definitive. Extracellular vesicles may also represent a promising minimally invasive biomarker source as they can carry cell-state-specific proteins, RNAs, and other molecular cargo reflective of underlying neurodegeneration and cell-specific pathological processes, however, current studies are preliminary and scarce [4]. Consequently, there is a critical unmet need for accessible, non-invasive biomarkers that reflect underlying FTD pathology [5].

The retina has emerged as a promising window into neurodegeneration, given its embryologic and structural continuity with the central nervous system [6]. Retinal layers can be quantitatively assessed using optical coherence tomography (OCT) [7, 8], offering a non-invasive method to detect parenchymal central nervous system loss [9]. Prior studies have suggested alterations in retinal structure in neurodegenerative diseases, including FTD, although findings have been inconsistent and limited by small sample sizes or a lack of eligible studies [10].

To address this, we conducted a systematic review and random-effects meta-analysis of OCT-derived retinal biomarkers for FTD. Our analysis suggests that individuals with FTD have significantly thinner retinal nerve fiber layer (RNFL) compared to AD, and significantly thinner ganglion cell layer-inner plexiform layer (GCL-IPL) compared to controls. However, more studies are needed to confirm these findings.

## Methodology

We conducted a systematic review, meta-analysis and meta-regression according to the protocol recommended by the Preferred Reporting Items for Systematic Reviews and Meta-Analyses Protocols (PRISMA). Our study only involved anonymized data, and we did not collect any personal information or perform any procedures on human subjects. Therefore, ethical approval was not necessary. We did not register the study’s protocol.

### Data sources and search strategy

We performed a thorough search for relevant articles by using specific search terms related to FTD. The search was conducted in two databases (PUBMED and EMBASE) and covered articles published from the inception of the databases until April 25th, 2026. The complete search strategy can be found in **Table S1**.

### Eligibility criteria

Eligible studies were required to assess retinal structural or vascular measures using optical coherence tomography (OCT) or OCT angiography (OCTA) at baseline in individuals with FTD alongside at least one comparator group, including Alzheimer’s disease (AD), other neurodegenerative disorders, or controls. We excluded studies involving animal models or cell lines, studies that did not include FTD or a comparator group, and studies that did not report sample size. A study with longitudinal measurements and follow-up was qualitatively discussed to ensure we did not duplicate individuals in the meta-analysis. Additionally, a study that evaluated individuals prior to phenoconversion was only discussed qualitatively because we wanted to examine diagnostic biomarker differences between FTD and AD or FTD and controls.

### Data extraction

Data from eligible studies were extracted by two independent researchers (EW and AK) and checked by a third researcher (HBT). We only performed meta-analyses on biomarkers that were measured by at least two different studies. The standard error of the mean for RNFL, GCL-IPL, and SCT thicknesses, and CVI was converted to standard deviation where required by multiplying the standard error of the mean by the square root of the number of eyes evaluated. Three studies presented median MMSE values, which we approximated as the mean through a previously described method by summing the Q1, median, and Q3, and dividing that sum by 3.

### Risk of bias assessment

Risk of bias was assessed using a modified Newcastle–Ottawa Scale for cross-sectional studies. Studies were categorized according to the context in which OCT or OCT-A was evaluated, rather than the original purpose of cohort recruitment. Risk of bias assessment was conducted by EW & AK and further checked by HBT.

### Data synthesis and statistics

If authors in the original study presented both adjusted and unadjusted values, the unadjusted values were used for meta-analyses. Meta-analyses were performed in R software (version 4.6.0). Pooled standardized mean differences and 95% CIs were calculated based on Cohen’s d. Funnel plots, Begg’s rank correlation, and Egger’s regression tests were used to evaluate publication bias [18]. Meta-analyses were based on a random effects model with inverse-sample weighting. Meta-regressions were used to investigate the association between OCT biomarkers with age and female percent.

## Results

We performed a systematic search (**Figure 1**) and included 10 [11–16, 19–22] studies in the systematic review, 5 of which we were able to meta-analyze. In our analysis, studies that grouped individuals as predicted or probable frontotemporal lobar degeneration tauopathy (pFTLD-Tau) were combined with FTD, and predicted or probable AD neuropathologic change were combined with AD.

**Figure 1.**
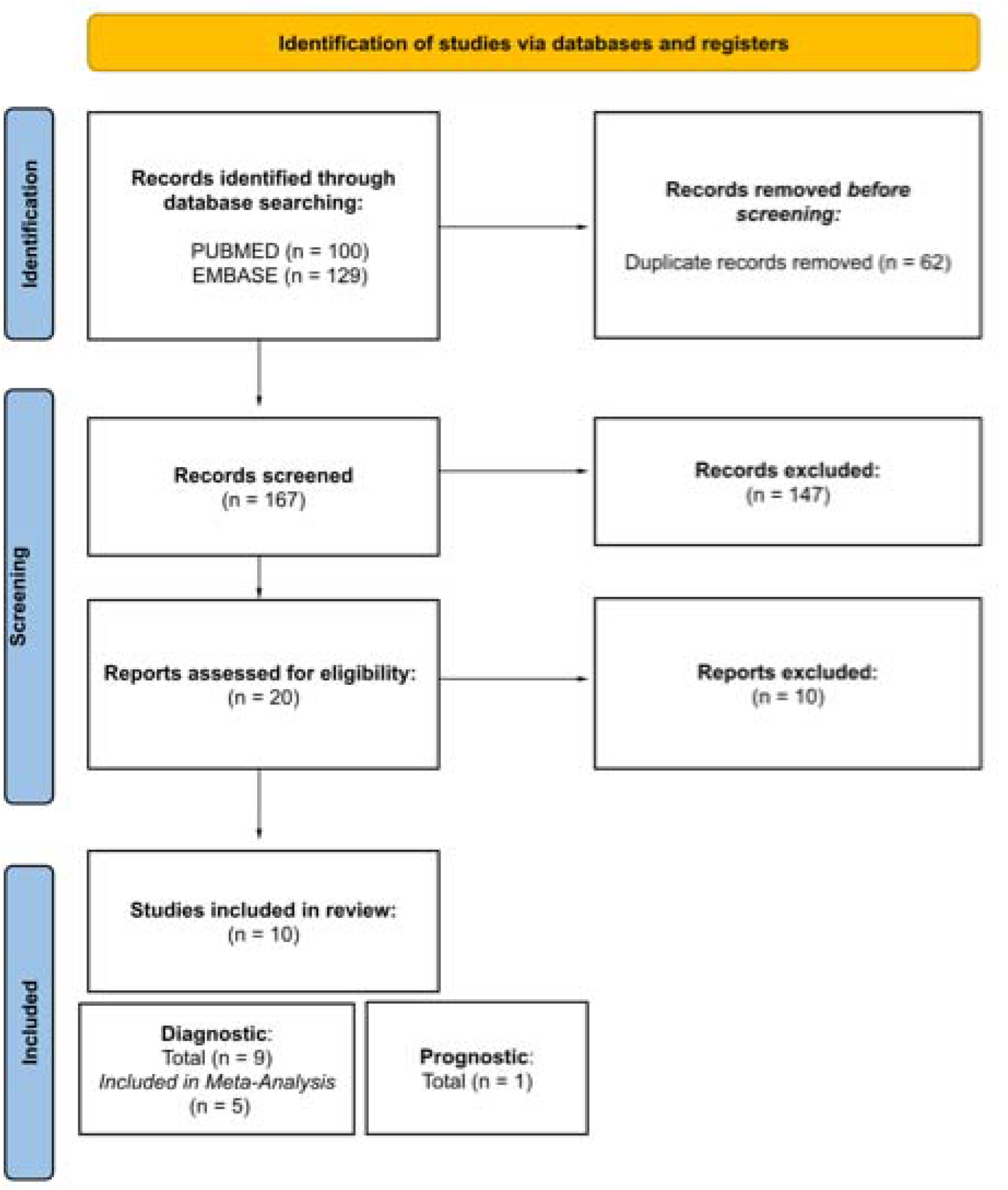
PRISMA flow-diagram.

The systematic review evaluated 139 individuals with FTD (55.4% female), 87 individuals with AD (52.9% female), 29 individuals with MCI (51.7% female), 14 individuals with TDP-43 proteinopathy (28.6% female), 5 individuals with tauopathy (60% female), and 255 controls (56.9% female). A total of 212 eyes from FTD participants, 132 eyes from AD participants, 29 eyes from MCI participants, 28 eyes from TDP-43 proteinopathy, 9 eyes from tauopathy, and 387 eyes from controls were assessed. The weighted mean age was 65.1 years in FTD, 65.3 years in AD, 70.5 years in MCI, 62.1 years in TDP-43 proteinopathy, 72.7 years in tauopathy, and 62.6 years in controls. Of the studies that presented MMSE scores, MMSE scores were 22.8 in FTD, 20.6 in AD, 26.6 in MCI, and 29.76 in controls.

One study reported that the number of eyes analyzed varied depending on the OCT/OCTA parameter measured. In all cases, the values reported in the tables reflect participant characteristics at baseline, except for one study that reported values reflecting the status of the original cohort at follow-up. Another study reported participant status prior to phenoconversion, thus, for our descriptive statistics, we excluded all cohorts from this study. Additionally, we note that 13 individuals with FTD from a previous study were reincluded in a subsequent study. The descriptive statistics and weighted means presented here count unique individuals, so we excluded the specific cohorts in studies that consisted entirely or partially of repeat individuals from our calculations.

Overall, included studies demonstrated relatively low risk of bias, with total Newcastle– Ottawa Scale scores ranging from 6/8 to 8/8 (**Table S2**). All studies were considered representative of the target population, used validated diagnostic or imaging assessment methods, and investigated potential confounders. Similarly, all studies relied on physician-based diagnostic criteria or established records for outcome ascertainment. The main sources of potential bias arose from limited sample sizes (<50 participants across groups in several studies) and insufficient reporting of response or attrition rates, with most studies receiving no score in the non-response domain. Only two studies achieved full scores for response rate reporting. These findings suggest generally good methodological quality, although small cohorts and incomplete reporting remain important limitations.

### FTD vs. AD

To explore whether OCT eye biomarkers differed between FTD and AD, we performed a random-effects meta-analysis on two studies. We found that FTD had a significantly thinner RNFL when compared to AD (SMD = −0.61, 95% CI: −0.98, −0.24]; **Figure 2A**). We did not find evidence of publication bias through funnel plot assessment (**Figure 2B**) or statistical Begg’s rank correlation and Egger’s tests (**Table S3-S4**).

**Figure 2.**
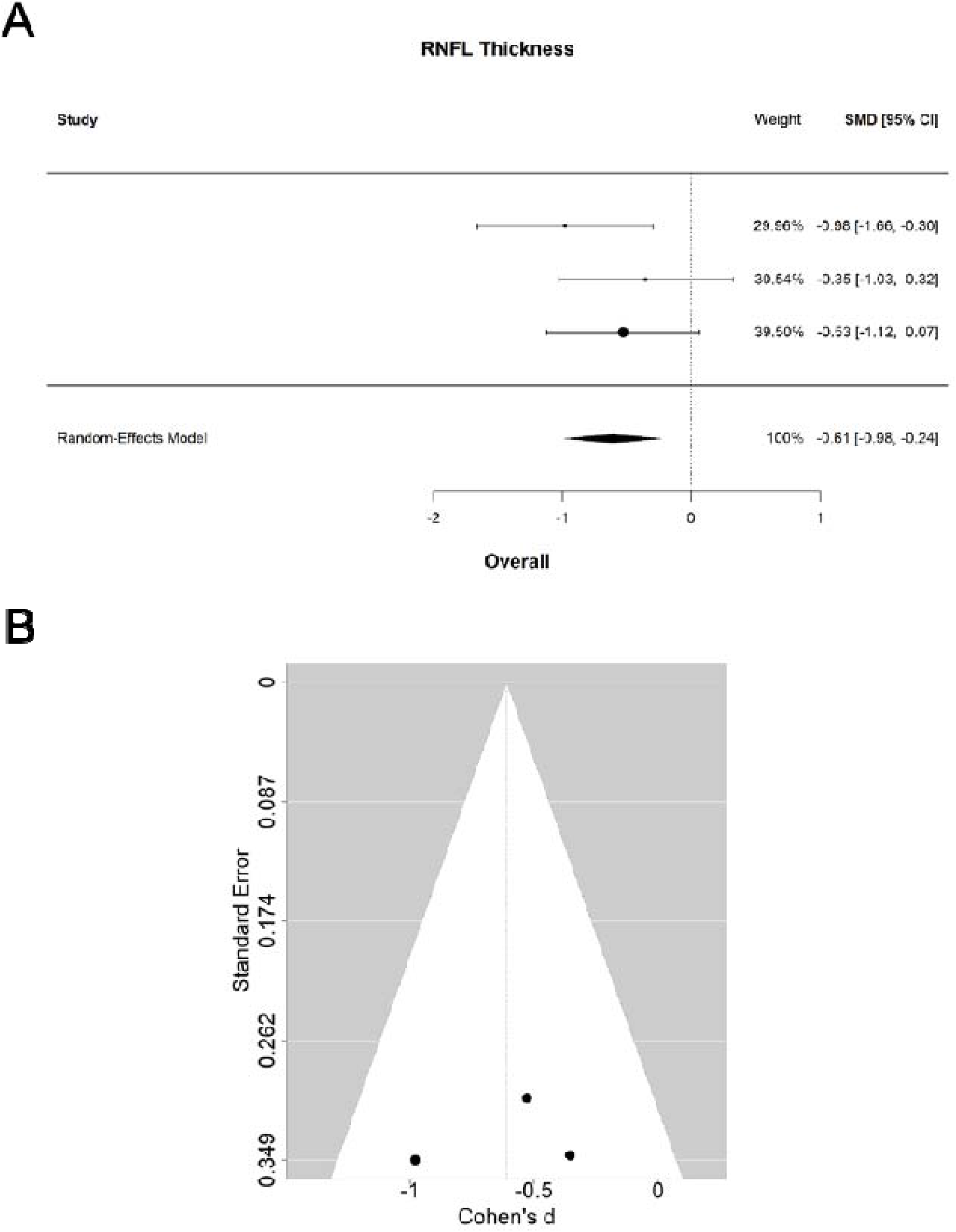
Forest and funnel plots evaluating retinal biomarker thickness differences between individuals with FTD or AD. Forest plot demonstrating standardized mean differences for RNFL thickness measurements comparing FTD and AD participants using an inverse variance random effects model (A). Funnel plot analysis evaluating publication bias and small study effects across included retinal biomarker studies (B). *Abbreviations:* AD, Alzheimer disease; pADNC, probable Alzheimer’s disease neuropathologic change; FTD, frontotemporal dementia; pFTLD-Tau, probable frontotemporal lobar degeneration with tauopathy; RNFL, retinal nerve fiber layer; SMD, standardized mean difference.

### FTD vs. Controls

To explore whether OCT eye biomarkers differed between FTD and controls, we performed a random-effects meta-analysis on four studies. We found that FTD had only a significantly thinner GCL-IPL compared to controls (SMD = −0.55, 95% CI: −1.02, −0.08; **Figure 3A**). When combined with other OCT biomarkers, the model became non-significant (SMD = −0.19, 95% CI: −0.52, 0.14; **Figure 3A**). We did not find evidence of publication bias through funnel plot assessment (**Figure 3B**) or statistical Begg’s rank correlation and Egger’s tests (**Table S3-S4**).

**Figure 3.**
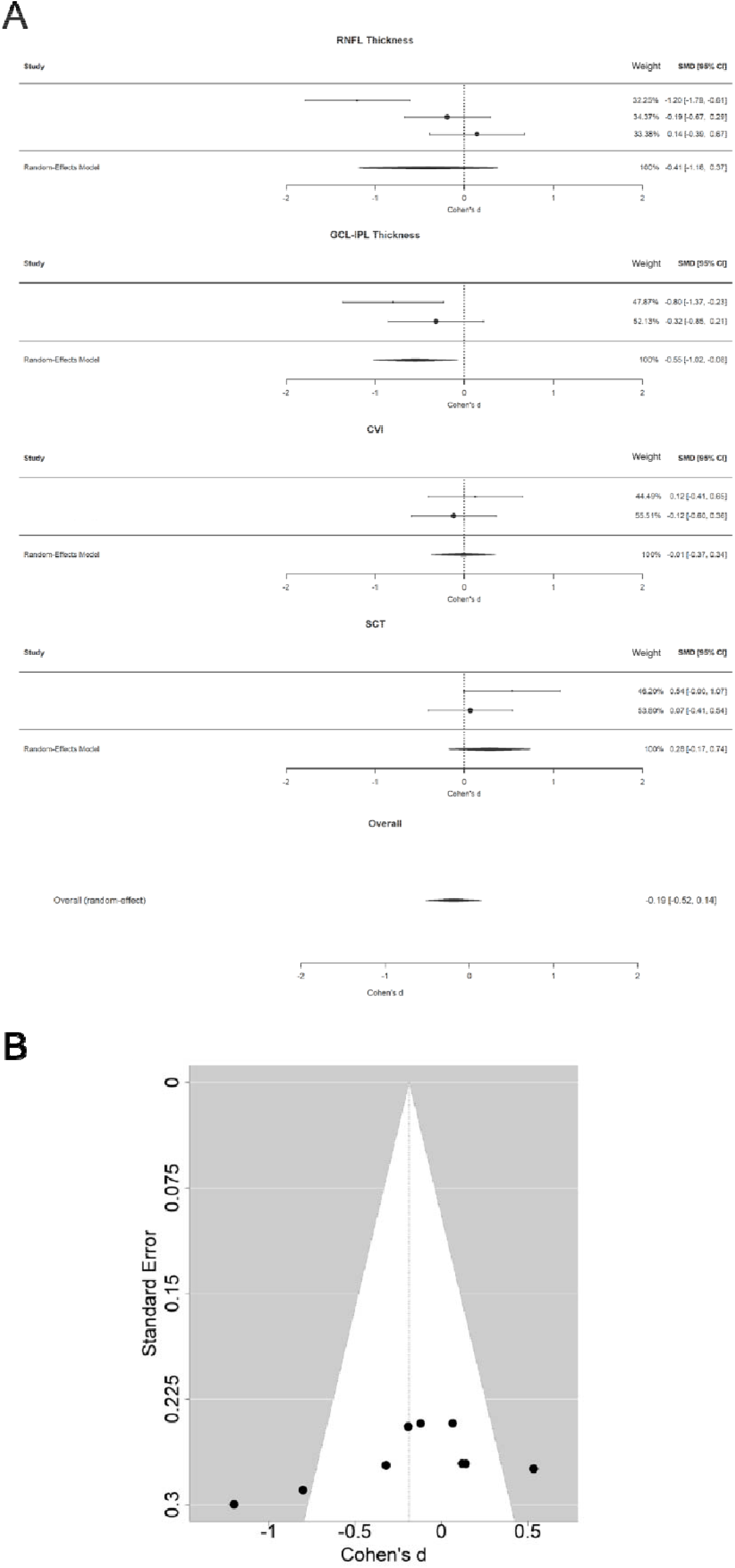
Forest and funnel plots evaluating retinal biomarker thickness differences between individuals with FTD and controls. Forest plots demonstrate standardized mean differences for RNFL and GCL-IPL thickness measurements, CVI, and SCT, comparing FTD and control participants using an inverse weighing random effects model (A). Funnel plot analysis evaluating publication bias and small study effects across included retinal biomarker studies (B). *Abbreviations:* FTD, frontotemporal dementia; pFTLD-Tau, predicted frontotemporal lobar degeneration with tauopathy; GCL-IPL, ganglion cell layer-inner plexiform layer; RNFL, retinal nerve fiber layer; CVI, choroidal vascularity index; SCT, subfoveal choroidal thickness; SMD, standardized mean difference.

### Meta-regression

To examine the impact of study-level covariates, we performed meta regressions to explore how biomarkers varied with patient demographics (age and female percent). We did not identify any significant correlations in the overall model for FTD (**Figure 4A-B**) and control (**Figure 4E-F**). Due to the large differences in scale of the raw measurement among each biomarker, we decided to perform meta regressions on individual biomarkers. When we specifically examined RNFL thickness, we found that RNFL thickness correlated negatively with female % in FTD and positively with age in both AD (**Figure 4C**) and control (**Figure 4E**), but there was no significant relationship between RNFL thickness and female percent for AD or controls (**Figure 4D, 4F)**. Given that CVI, GCL-IPL, and SCT each only had two data points, we did not correlate them with age and female %, but close inspection of the plots suggests minimal correlation. Meta-regressions statistics are summarized in **Table S5**.

**Figure 4.**
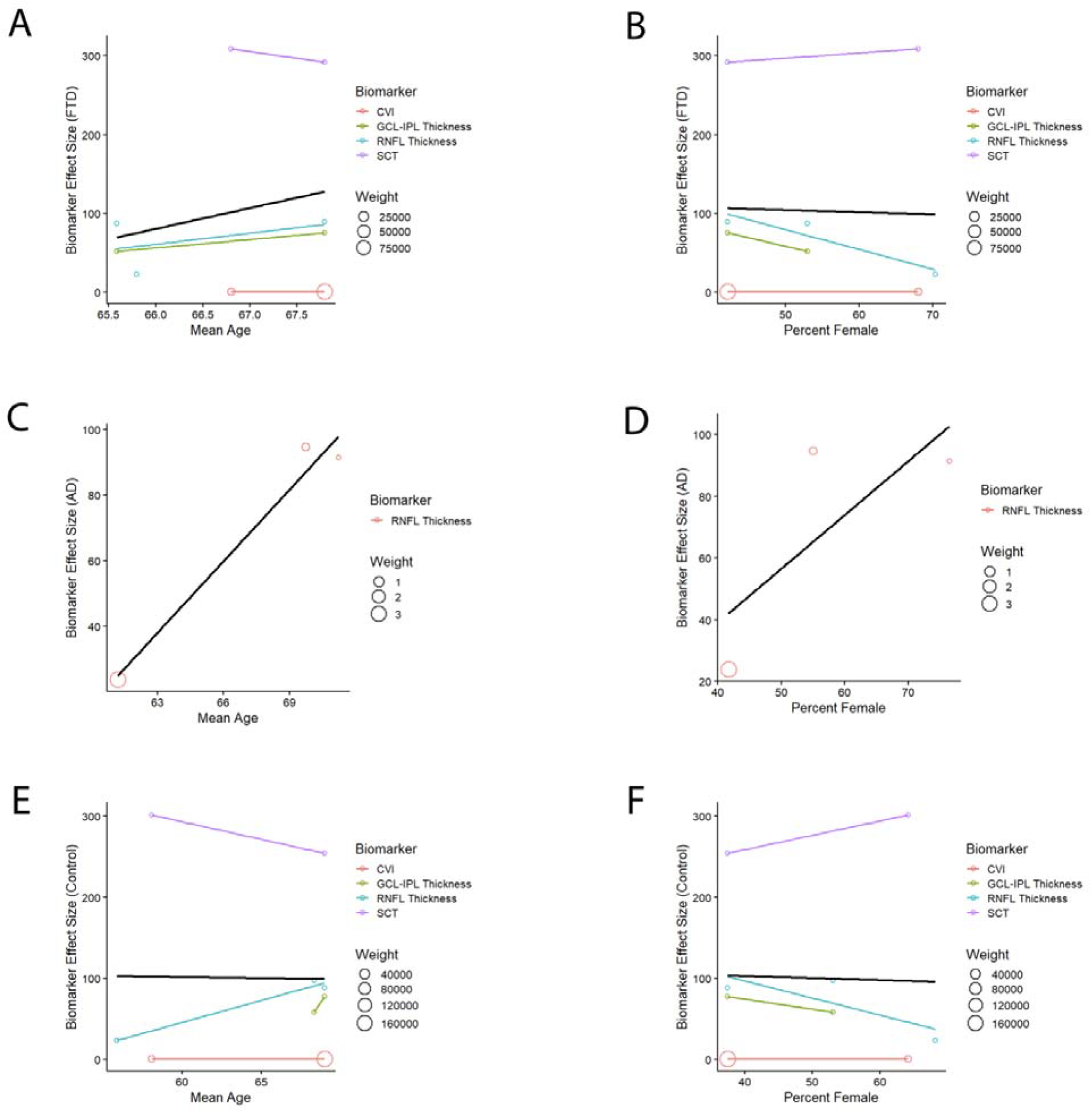
Meta regression analyses evaluating associations between demographic characteristics and retinal biomarker effect sizes across FTD, AD, and controls. Associations between retinal biomarker effect size and mean age are shown for FTD (A), AD (C), and controls (E), while associations between retinal biomarker effect size and percent female participants are shown for FTD (B), AD (D), and controls (F). Bubble size reflects study weight, and black regression lines represent overall meta regression trends across included biomarkers. *Abbreviations:* AD, Alzheimer disease; FTD, frontotemporal dementia; GCL-IPL, ganglion cell layer-inner plexiform layer; RNFL, retinal nerve fiber layer; CVI, choroidal vascularity index; SCT, subfoveal choroidal thickness.

Additionally, two studies performed cross-sectional correlations between OCT/OCT-A biomarkers and measures of disease severity. One study found a positive correlation between the choroidal vascularity index and disease duration only in pADNC, but not with choroidal thickness measurements and disease duration in either pFTLD-Tau or pADNC. Another study found that GCL-IPL thickness correlated positively with cognitive dysfunction scores (MMSE scores) only in persons with AD, while RNFL thickness did not correlate with MMSE in FTD, AD or controls. Adjusting for age, sex and disease duration did not attenuate the GCL-IPL thickness and cognitive dysfunction score association in AD. In a longitudinal study comparing individuals with FTD to controls, both the follow-up OCT measurements for ONL and outer retina thicknesses and change in ONL and outer retina thicknesses had a positive correlation with follow-up MMSE scores. Furthermore, the change in outer retina thickness as measured in the probable tauopathy subgroup of individuals with FTD was positively correlated with follow-up MMSE scores.

## Discussion

Current antemortem biomarkers for accurate diagnosis of FTD from other dementias or controls are lacking. Changes in the retina may serve as a non-invasive window to biochemical changes during neurodegeneration given its embryologic and structural continuity with the central nervous system. In this study, we included 10 studies in the systematic review, 5 of which we meta-analyzed. In the meta-analysis, we found that individuals with FTD had lower RNFL thickness compared to AD (**Figure 2A**) and lower GCL-IPL thickness compared to controls (**Figure 3A**) with no publication bias (**Figure 2B, 3B, Table S3-S4**). Meta-regression analyses did not identify significant associations between retinal biomarker effect sizes and age in FTD; however, RNFL thickness was significantly associated with female % in FTD, while RNFL thickness correlated positively with age in AD and control groups (**Figure 4, Table S5**).

In several studies inner retinal layers (e.g. RNFL and GCL) in individuals with FTD and AD did not differ from controls. However, the outer retina, outer nuclear layer (ONL), and ellipsoid zone were thinner in FTD compared to controls. Upon follow up 1-2 years later, there was progressive outer retinal thinning compared to baseline in the tauopathy subgroup of FTD with no corresponding change in inner retinal layers. In another study, when comparing FTD and AD, the ONL was thinner in individuals with pFTLD-Tau than individuals with pADNC but ONL correlated with worse MMSE and higher CDR-FTLD sum of boxes scores only in pFTLD-Tau and not pADNC. The nuances in OCT biomarkers are further underscored by how ganglion cell-inner plexiform layer (GC-IPL) thinning was associated with increased AD risk, but thinning of the retinal pigment epithelium in the inner superior subfield was linked to increased FTD risk. When specifically comparing between TDP-43 and tauopathy pathologies, TDP-43 had a thinner peripapillary RNFL which was not significant after adjustment for multiple comparisons.

Three studies used OCT-A to investigate vascular eye biomarkers for individuals with FTD compared to controls. Compared to controls, FTD had reduced 3×3mm macular perfusion density, vessel density, and capillary flux index. Individuals with FTD also had reduced optic nerve head and macular superficial capillary plexus flow density, as well as reduced vascular density in the macular deep capillary plexus compared to controls; these changes did not correlate with brain MRI data, cognitive deficits, or cerebrospinal fluid markers. In contrast, reduced perfusion to the retina and choriocapillaris was observed in individuals with FTD compared to controls. When MRI data was analyzed, the decrease in superficial vascular complex density was found to be correlated with elevated white matter hypersensitivity and perivascular space volumes.

Several studies have also conducted ROC analyses to distinguish FTD from AD or FTD from controls. In one study, ROC analysis differentiating FTD from controls demonstrated an AUC of 0.72 (95% CI 0.60–0.84) for outer retina thickness alone, which improved to 0.81 (95% CI 0.71–0.91) when combined with age. Another study distinguishing pFTLD-Tau from pADNC using ONL thickness reported an AUC of 0.78 (95% CI 0.64–0.89), which further improved to 0.85 (95% CI 0.72–0.94) following adjustment for MMSE scores. Additionally, OCTA metrics demonstrated strong discriminatory performance between FTD and controls, with reported AUCs ranging from 0.76 to 0.99 across retinal vascular layers and choriocapillaris measurements. We were not able to conduct a diagnostic accuracy meta-analysis due to missing statistics.

We note several limitations in this study. First, although 10 studies were included in the systematic review, only 5 studies were eligible for meta-analysis, limiting statistical power. Second, there is potential selection bias, as 4 of the 10 included studies originated from the same research group and some participants were reincluded across studies. Third, there was substantial heterogeneity in OCT/OCTA methods, retinal biomarkers measured, diagnostic group definitions, and statistical adjustments across studies. Fourth, many studies had small sample sizes and cross-sectional designs with limited longitudinal follow-up. Additionally, most studies relied on clinical rather than neuropathologic diagnoses of FTD subtypes, which may have introduced diagnostic misclassification. Future studies should include larger multicenter cohorts, standardized OCT/OCTA protocols, and longitudinal follow-up to better determine how retinal biomarkers change over time and whether they can distinguish FTD subtypes or predict disease progression.

## Conclusion

Individuals with FTD had lower RNFL thickness compared to AD and lower GCL-IPL thickness compared to controls. However, future studies with diverse and larger longitudinal cohorts and standardized OCT/OCTA methodologies are needed to better determine the diagnostic and prognostic utility of retinal biomarkers in FTD.

## Data Availability

Data is available upon request.

## Acknowledgment

None

