## Supplementary material for "Optical coherence tomography as a biomarker for frontotemporal dementia: a systematic review & meta-analysis": Table S1

**Table S1.** **Complete Search Strategy using PUBMED and EMBASE. The search was performed from the date of inception until April 25th, 2026.**

| PUBMED | ("Frontotemporal Dementia"[Mesh] OR "frontotemporal dementia"[tw] OR FTD[tw] OR FTLD[tw] OR "frontotemporal lobar degeneration"[tw] OR frontotemp*[tw])  AND  ("Tomography, Optical Coherence"[Mesh] OR "optical coherence tomography"[tw] OR OCT[tw] OR OCTA[tw] OR "optical coherence tomography angiography"[tw] OR retina*[tw] OR fundus[tw] OR "retinal nerve fiber layer"[tw] OR RNFL[tw] OR "ganglion cell layer"[tw] OR GCL[tw] OR "retinal thickness"[tw] OR macula*[tw] OR choroid*[tw] OR peripapillary[tw])  NOT  ("systematic review"[pt] OR "review"[pt] OR "case reports"[pt] OR "clinical conference"[pt] OR "editorial"[pt] OR "meta-analysis"[pt])  NOT  (animals[Mesh] NOT humans[Mesh]) |
| --- | --- |
| EMBASE | ('frontotemporal dementia'/exp OR 'frontotemporal dementia':ti,ab,kw OR ftd:ti,ab,kw OR ftld:ti,ab,kw OR 'frontotemporal lobar degeneration':ti,ab,kw OR frontotemp*:ti,ab,kw) AND ('optical coherence tomography'/exp OR 'optical coherence tomography':ti,ab,kw OR oct:ti,ab,kw OR octa:ti,ab,kw OR 'optical coherence tomography angiography':ti,ab,kw OR retina:ti,ab,kw OR retinal:ti,ab,kw OR fundus:ti,ab,kw OR 'retinal nerve fiber layer':ti,ab,kw OR rnfl:ti,ab,kw OR 'ganglion cell layer':ti,ab,kw OR gcl:ti,ab,kw OR 'retinal thickness':ti,ab,kw OR macula*:ti,ab,kw OR choroid*:ti,ab,kw OR peripapillary:ti,ab,kw) NOT ('systematic review'/de OR 'review'/de OR 'meta analysis'/de OR 'case report'/de OR 'conference abstract':it OR 'editorial':it) NOT ('animal'/exp NOT 'human'/exp) |
